# Deep Learning for Heart Sound Analysis: A Literature Review

**DOI:** 10.1101/2023.09.16.23295653

**Authors:** Qinghao Zhao, Shijia Geng, Boya Wang, Yutong Sun, Wenchang Nie, Baochen Bai, Chao Yu, Feng Zhang, Gongzheng Tang, Deyun Zhang, Yuxi Zhou, Jian Liu, Shenda Hong

**Affiliations:** Department of Cardiology, Peking University People’s Hospital, Beijing, China; HeartVoice Medical Technology, Hefei, China; Key laboratory of Carcinogenesis and Translational Research (Ministry of Education/Beijing), Department of Gastrointestinal Oncology, Peking University Cancer Hospital and Institute, Beijing, China; Department of Computer Science, Tianjin University of Technology, Tianjin, China; DCST, BNRist, RIIT, Institute of Internet Industry, Tsinghua University, Beijing, China; National Institute of Health Data Science, Peking University, Beijing, China; Institute of Medical Technology, Health Science Center of Peking University, Beijing, China

**Keywords:** heart sounds, phonocardiogram, artificial intelligence, deep learning

## Abstract

Heart sound auscultation is a physical examination routinely used in clinical practice to identify potential cardiac abnormalities. However, accurate interpretation of heart sounds requires specialized training and experience, thereby limiting its generalizability. Deep learning, a subset of machine learning, involves training artificial neural networks to learn from large datasets and perform complex tasks related to intricate patterns, such as disease diagnosis, event prediction, and clinical decision-making. Over the past decade, deep learning has been successfully applied to heart sound analysis with remarkable achievements. Meanwhile, as heart sound analysis is gaining attention, many public and private heart sound datasets have been established for model training. The massive accumulation of heart sound data improves the performance of deep learning-based heart sound models and extends their clinical application scenarios. In this review, we will compile the commonly used datasets in heart sound analysis, introduce the fundamentals and state-of-the-art techniques in heart sound analysis and deep learning, and summarize the current applications of deep learning for heart sound analysis and their limitations for future improvement.

## 1 Introduction

Heart sounds are generated from the turbulent movement of blood through the heart chambers as the valves open and close during the cardiac cycle, which can be affected by anatomical changes or narrow blood vessels [1]. Cardiac auscultation, which involves listening to heart sounds with a stethoscope, is a non-invasive and easy-to-operate technique widely accepted by medical professionals to identify potential cardiac abnormalities. However, the interpretation of heart sounds can vary greatly depending on the experience and skill of the examiners. Even in detecting the systolic murmurs, which is a common task in auscultation, the reliability is mediocre at best (*k* = 0.3 − 0.48) [2], and the ability to identify other pathological features is even worse [3]. In addition, the growing availability of recording techniques has led to a substantial accumulation of heart sound data, particularly long-term heart sound monitoring. However, manual analysis using traditional auscultation methods is no longer sufficient to handle the data surge, especially in real-time scenarios.

To address these challenges, researchers are actively developing computational methods for analyzing heart sound data, whose graphical representation is known as phonocardiogram (PCG). In recent years, the potential of deep learning (DL) techniques in PCG analysis has gradually attracted the interest of researchers. DL is suitable for tasks involving a large amount of data with complex patterns, and the PCG signal has a high sampling frequency and contains rich information in both the time and frequency domains, consistent with this data characteristic. DL models can be trained using either raw signals in the time domain or pre-processed frequency information, allowing them to capture subtle patterns in heart sounds that may be difficult for physicians to discern. By leveraging this underlying knowledge, these models have demonstrated remarkable diagnostic accuracy across a range of cardiac conditions [4, 5]. Furthermore, DL-based heart sound diagnostic techniques offer a cost-effective and user-friendly approach, making them a more economical and accessible alternative for disease screening than traditional medical imaging techniques. This advantage is precious for undiagnosed patients in underdeveloped regions who may face limited access to specialized medical imaging equipment and trained healthcare providers.

In this review, we will introduce the datasets commonly used in heart sound analysis, as these datasets play a fundamental role in advancing research in this field. Subsequently, we will demonstrate the knowledge of heart sound analysis and DL, highlighting why DL is a suitable method for analyzing heart sounds. Then, we will comprehensively summarize the clinical applications of DL-based heart sound analysis. Finally, we will conclude with a summary of the current challenges and future perspectives in this field.

## 2 Methods

### 2.1 Search strategy

To summarize current research findings on heart sound analysis using DL, we searched PubMed, Embase, Web of Science, and Google Scholar for “deep learning” or “machine learning” or “artificial intelligence” in conjunction with “heart sounds” or “cardiac sounds” or “heart murmur” or “cardiac murmur” or “phonocardiogram”, “phonocardiography” or “PCG”, and covers the period from January 1st 2010 to January 1st 2023. All keywords are case-insensitive. To avoid missing papers that did not explicitly mention these keywords in their titles, we expanded our search to include all fields in each article. In total, 211 related studies were found.

### 2.2 Study selection

We only included published peer-reviewed articles and excluded reviews, editorials, non-heart sound studies, non-artificial intelligence studies, and papers written in foreign languages. As this review focused on DL, we excluded those studies with a narrow sense of machine learning involved with conventional machine learning algorithms and applications outside the DL regime. However, we retained research on neural networks and their variants because of their close proximity to DL model structures.

The process of literature searching and selection is illustrated in Figure 1. At last, 71 original articles were included. These studies can be broadly categorized into several groups: methods (15 papers, including heart sound segmentation [6, 7, 8, 9, 10, 11, 12, 13], noise cancellation [14, 15, 16], algorithm development [17, 18, 19], and database development [20]), cardiac murmurs detection (36 papers [21, 22, 23, 24, 25, 26, 27, 28, 29, 30, 31, 32, 33, 34, 35, 36, 37, 38, 39, 40, 41, 42, 43, 44, 45, 46, 47, 48, 49, 50, 51, 52, 53, 54, 55, 56]), valvular heart disease (6 papers [57, 58, 59, 60, 61, 62]), congenital heart disease (4 papers [63, 64, 65, 66]), heart failure (4 papers [67, 68, 69, 70]), coronary artery disease (2 papers [71, 72]), rheumatic heart disease (2 papers [73, 74]), and extracardiac applications (2 papers [75, 76]).

**Figure 1:**
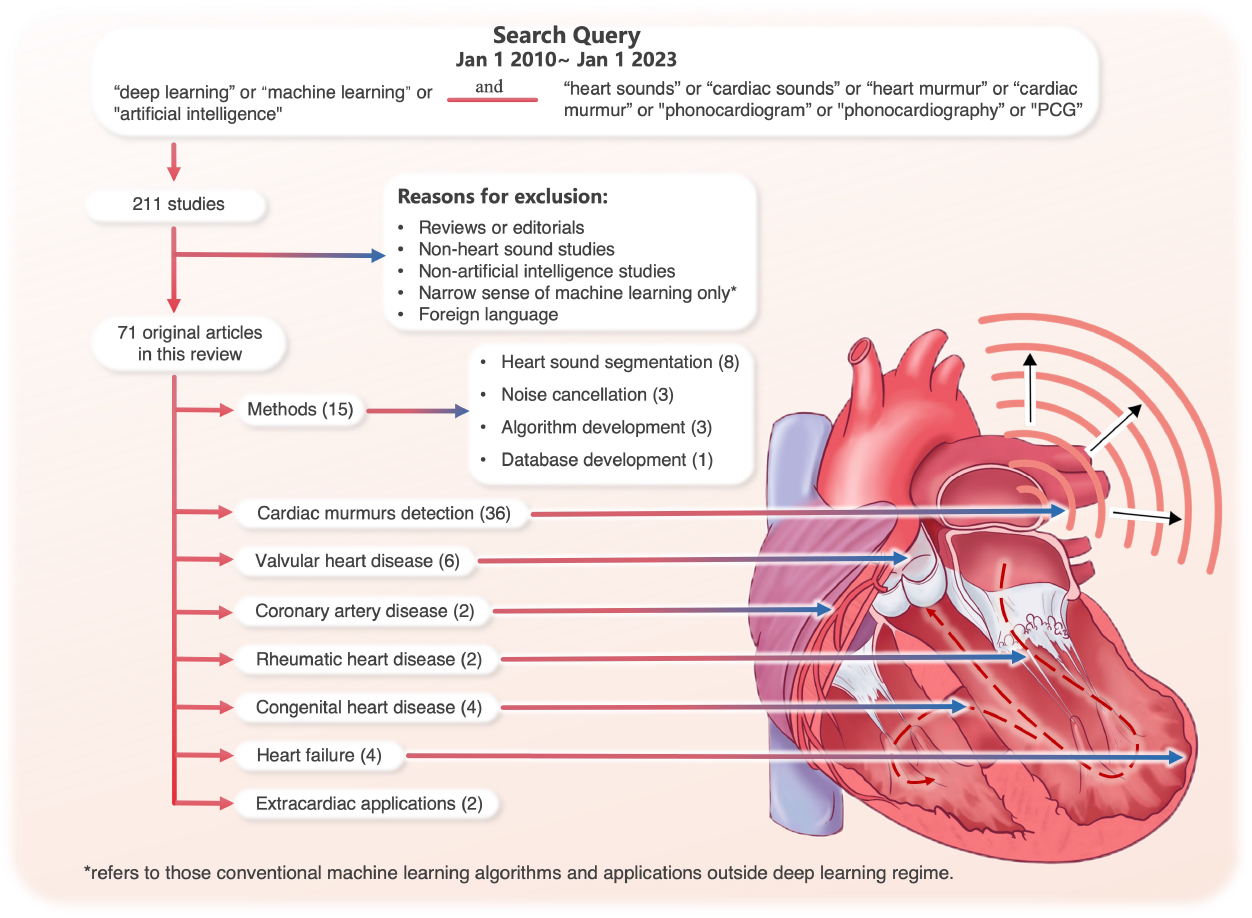
Diagram demonstrating literature selection process and applications of deep learning in heart sound analysis

## 3 Results

### 3.1 Heart sound datasets

The study of sound is a discipline with ancient roots that can be traced back centuries. Before the invention of modern medical imaging techniques, physicians heavily relied on sounds to gain insights into the inner workings of the body, particularly heart sound auscultation. As recording technology has advanced, it has become possible to capture heart sounds in both analog and electronic forms, leading to the accumulation of vast amounts of data that can be utilized for in-depth analysis. This has facilitated the availability of publicly-accessible datasets that serve as valuable resources for benchmarking and testing novel methods and approaches in heart sound analysis.

Table 1 showcases a selection of well-known public heart sound datasets. The HSS dataset [77] is unique in detailing the severity levels of heart conditions. PASCAL [78], CinC2016 [79], and CinC2022 [20] are suitable for distinguishing between normal and abnormal heart conditions. On the other hand, the Khan dataset [80] focuses on specific valvular heart diseases. The EPHNOGRAM dataset [81] stands out for providing simultaneous electrocardiogram and heart sound recordings during fitness exercises. Additionally, the SUFHSDB dataset [82] offers fetal and maternal heart sounds.

**Table 1:** Public heart sound datasets.

| Dataset | Resource | Sampling Frequency | Recording Duration | Number of Recordings and/or Subjects (if mentioned) |
| --- | --- | --- | --- | --- |
| HSS [77] | patients with various health conditions from the hospital | 4,000 Hz | 30 s on average | 845 recordings (144 normal, 465 mild, 236 moderate/severe) from 170 subjects |
| PASCAL [78] | two sources: (A) from the general public via the web and (B) from hospital patients | (A) 44,100 Hz;<br>(B) 4,000 Hz | 1 s to 30 s | (A) 124 recordings (31 normal, 34 murmur, 19 extras, 40 artifacts)<br>(B) 461 recordings (320 normal, 95 murmur, 46 artifacts) |
| Khan [80] | books (Auscultation skills CD, Heart sound made easy) and 48 websites | 8,000 Hz | 3 s to 5 s | 1,000 recordings (200 for each: normal, MVP, MR, MS, AS) |
| CinC2016 [79] | nine databases from different research groups | 2,000 Hz | 5 s to 120 s | 3,240 recordings (2,575 normal, 665 abnormal with CAD or VHD.) |
| CinC2022 [20] | two mass pediatric screening campaigns conducted in Brazil | 4,000 Hz | 4.8 s to 80.4 s | 5,272 recordings from 1,568 subjects<br>(486 normal subjects, 1,082 subjects with heart diseases) |
| EPHNOGRAM [81] | healthy adults indoor fitness, simultaneous electrocardiogram and phonocardiogram | 8,000 Hz | 30 s or 30 min | 69 recordings (10 bicycle stress test, 11 treadmill, 13 static bike, 35 resistance training) from 24 subjects |
| SUFHSDB [82] | fetal and maternal heart sounds obtained at Hafez Hospital, Shiraz University | 16,000 Hz; 44,100 Hz | 90 s on average | 119 recordings from 109 subjects |

In addition to publicly available datasets, numerous researchers collected their own heart sound data for specific research purposes. Table 2 provides a comprehensive overview of various private heart sound datasets and their applications. These datasets originate from diverse sources and cover various heart-related conditions, such as valvular heart diseases, heart failure, and congenital heart diseases. These datasets exhibit diversity in sampling frequency, spanning from 2,000 Hz to 8,000 Hz, and provide recordings of varying durations, ranging from short 5-second clips to extensive 120-second recordings. Their scale also varies significantly, from only dozens to thousands of subjects. Each dataset has been cited in one or more research studies, emphasizing their importance in advancing the understanding and analysis of heart sounds.

**Table 2:** Private heart sound datasets.

| Dataset | Application | Resource | Sampling Frequency | Recording Duration | Number of Recordings and/or Subjects (if mentioned) |
| --- | --- | --- | --- | --- | --- |
| Private heart murmur dataset<br>Chorba et al. (2021) [61] | Heart murmur detection | patients from four echocardiography laboratories and structural heart disease clinics in the US | 4,000 Hz | 15 s × 4 positions | 1,774 recordings (682 murmur, 1092 normal) from 962 subjects |
| Private VHD dataset<br>Makimoto et al. (2022) [62] | Valvular heart disease | patients at the Faculty of Medicine at Heinrich Heine University Düsseldorf, Germany. | 4,000 Hz | 15 s × 3 positions | 836 subjects (670 normal, 51 mild AS, 51 moderate AS, 114 severe AS) |
| Private CHD dataset 1<br>Gharehbaghi et al. (2017) [63] | Congenital heart disease | children referrals to the Children Medical Centre Hospital of Tehran University, Iran. | - | 10 s | 90 subjects (30 VSD, 15 MR, 15 TR, 30 normal) |
| Private CHD dataset 2<br>Wang et al. (2020) [64] | Congenital heart disease | patients at the National Taiwan University Hospital | - | 10 s × 5 positions × 2 | 776 recordings (525 VSD, 251 normal) from 76 subjects |
| Private CHD dataset 3<br>Liu et al. (2022) [65] | Congenital heart disease | children admitted to Children's Hospital of Chongqing Medical University, China | 22,050 Hz | 10 s × 5 positions | 884 subjects (409 normal, 192 ASD, 98 VSD, 95 PDA, 90 Combined CHD) |
| Private CHD dataset 4<br>Gharehbaghi et al. (2020) [66] | Congenital heart disease | children referrals to Tehran University of Medical Sciences, Iran | 44,100 Hz | 10 s | 115 subjects (10 ASD, 25 innocent murmur, 15 MR, 15 TR, 25 VSD, 25 normal) |
| Private HF dataset 1<br>Gao et al. (2020) [67] | Heart failure | patients at University-Town Hospital of Chongqing Medical University, China | 11,025 Hz | - | 108 subjects (42 HFrEF, 66 HFpEF) |
| Private HF dataset 2<br>Wang et al. (2022) [68] | Heart failure | patients at the First Affiliated Hospital of Chongqing Medical University, China | 4,000 Hz | 3 min | 136 subjects (59 HFrEF, 77 HFpEF) |
| Private HF dataset 3<br>Yang et al. (2021) [69] | Heart failure | patients at the First Affiliated Hospital of Chongqing Medical University, China | 8,000 Hz | 5 min | 71 subjects (30 LVDD, 41 normal) |
| Private HF dataset 4<br>Zheng et al. (2022) [70] | Heart failure | patients at the First Affiliated Hospital and the University-Town Hospital of Chongqing Medical University, China | 8,000 Hz | 5 min | 224 HF subjects (42 Stage A, 56 Stage B, 75 Stage C, 51 Stage D), 51 normal |
| Dan-NICAD trial 1 dataset<br>Winther et al. (2021) [83] | Coronary artery disease | patients from the Danish study of the Non-Invasive Testing in Coronary Artery Disease (Dan-NICAD) trial 1 | - | 3 min<br>(8 s holding breath × 4) | 1,464 subjects (723 CAD-score ≤ 20, 741 CAD-score $>$ 20) |
| Private CAD dataset<br>Li et al. (2021) [71];<br>Li et al. (2020) [72] | Coronary artery disease | patients at Shandong Provincial Qianfoshan Hospital, China | 1,000 Hz | 5 min | 195 subjects (135 CAD, 60 non-CAD) |
| Private RHD dataset<br>Asmare et al. (2020) [73] | Rheumatic heart disease | patients at Tikur Anbessa Referral Teaching Hospital, College of Health Sciences, Addis Ababa University, Ethiopia | 44,100 Hz | - | 170 subjects (124 RHD, 46 normal) |
| Proposed RHD dataset<br>Ali et al. (2021) [74] | Rheumatic heart disease | children from a group of schools serving the underprivileged in Karachi, Pakistan | - | - | aim to recruit 1,700 children (definite RHD, subclinical RHD, normal) |
| Private blood pressure dataset<br>Kapur et al. (2019) [75] | Blood pressure measurement | Critically ill children undergoing continuous blood pressure monitoring at the Children's Hospital of Michigan/Wayne State University, US | - | $>1\text{min} \times 2$ positions simultaneously | 25 subjects |

### 3.2 Heart sound analysis technologies

#### 3.2.1 Pre-processing and feature extraction

Because sound analysis has a long and well-established research history, heart sound analysis often draws upon the techniques from traditional sound studies. The input of DL models can be the features extracted using various time-frequency methods or a time representation that has undergone basic manipulations such as smoothing, denoising, and segmentation. The latter paradigm eliminates the need for a feature extraction step, allowing the model to directly process the raw data and output the task results—an “end-to-end” approach. Figure 2 illustrates the different process pathways.

**Figure 2:**
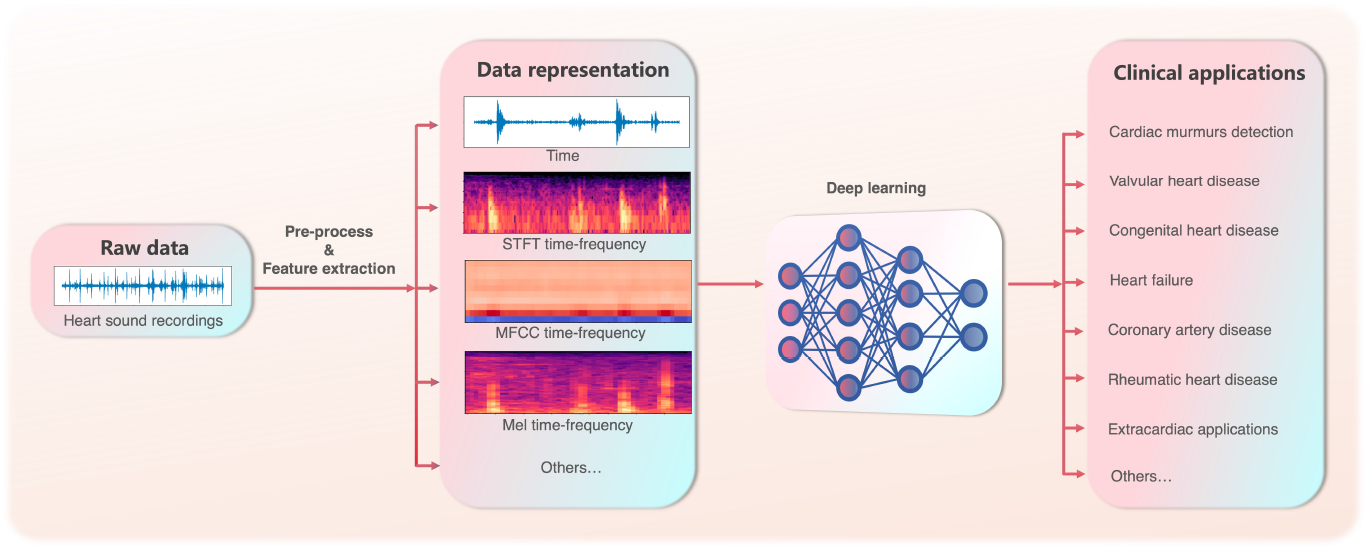
Different heart sounds process pathways.

#### 3.2.2 Deep learning

DL, a subset of machine learning (ML), involves training artificial neural networks to learn from large datasets and perform complex tasks related to human cognitive activities and experiences. It has been successfully applied to various tasks, including image classification, speech recognition, natural language processing, and disease diagnosis [85, 86, 87, 88, 89]. DL models perform intricate functions based on large numbers of simple non-linear computational units (known as artificial neurons) connected in complex hierarchical networks. This structure encourages each layer to learn simple representations that build up to sophisticated concepts. Compared to traditional ML, the fundamental architectural features of DL determine its greater ability to perform cohesive tasks, such as visual and computational knowledge representation. Another notable advantage of DL models is their ability to process raw data and automatically learn important features. Unlike traditional ML, which often requires handcrafted features, DL models can exploit some underlying features in raw data, enhancing classification accuracy and reducing the reliance on manual labeling.

The two most widely used DL model architectures are convolutional neural networks (CNNs) and recurrent neural networks (RNNs). CNNs are commonly used for grid-like data such as images and spectrograms. They employ multiple layers of convolutional filters to extract input features, followed by pooling layers to reduce the data dimensions. The output is then fed into fully connected layers for classification or regression tasks. RNNs, on the other hand, are designed to handle data with temporal dependencies, such as natural language and speech data. They process input sequences one unit at a time based on the current input unit and a hidden state that captures information from previous time steps.

In recent years, the Transformer architecture has gained significant popularity in DL. Initially developed for natural language processing, the Transformer has been adapted for various types of sequential input data, such as video, audio, and music [90, 91, 92]. Unlike RNNs, the Transformer can process the entire input sequence simultaneously using the attention mechanism, which allows it to focus on specific parts of the input sequence based on their relevance [93]. Consequently, the Transformer exhibits superior efficiency in both model training and inference stages.

Despite being one-dimensional signals, heart sounds pose challenges for traditional processing and analysis techniques due to their high sampling frequency and large number of samples per cardiac cycle. However, the emergence of DL models like CNNs, RNNs, and Transformers has made it possible to build high-performance models for heart sound analysis. These DL models can identify specific features relevant to various cardiac conditions, enhancing diagnostic accuracy and speed compared to manual methods. Moreover, DL techniques facilitate monitoring long-term changes in heart sounds, making them a powerful tool for continuous cardiac health tracking. By analyzing the evolving patterns and trends in heart sounds, healthcare professionals can detect and respond to cardiac abnormalities in a timely manner.

In the reviewed literature, most studies use CNN models with 2 to 34 convolutional layers [9, 10, 11, 13, 14, 15, 18, 21, 22, 23, 24, 25, 26, 27, 28, 31, 32, 33, 34, 35, 37, 38, 40, 41, 42, 47, 48, 50, 51, 52, 55, 58, 59, 60, 61, 68, 69, 71, 72, 73, 84], which are usually equipped with rectified linear units, batch normalization, dropout and pooling components, and some of the layers are linked by residual connections. Wang et al. [76] test 10 different CNN models including GoogleNet, SqueezeNet, DarkNet19, ModileNetv2, Inception-ResNetv2, DenseNet201, Inceptionv3, ResNet101, NasNet-Large, and Xception to compare the performances. Since heart sounds are continuous sequence signals, it is also suitable to test RNN models [8, 12, 49, 66], while other studies combine both CNN and RNN [29, 64, 65, 67, 74]. Other models include Transformer [7], traditional neural network (NN), and some NN modifications such as time delay neural network, time growing neural network, and kernel sparse representation network. Table 3 lists the heart sounds analysis models in the reviewed articles.

**Table 3:**
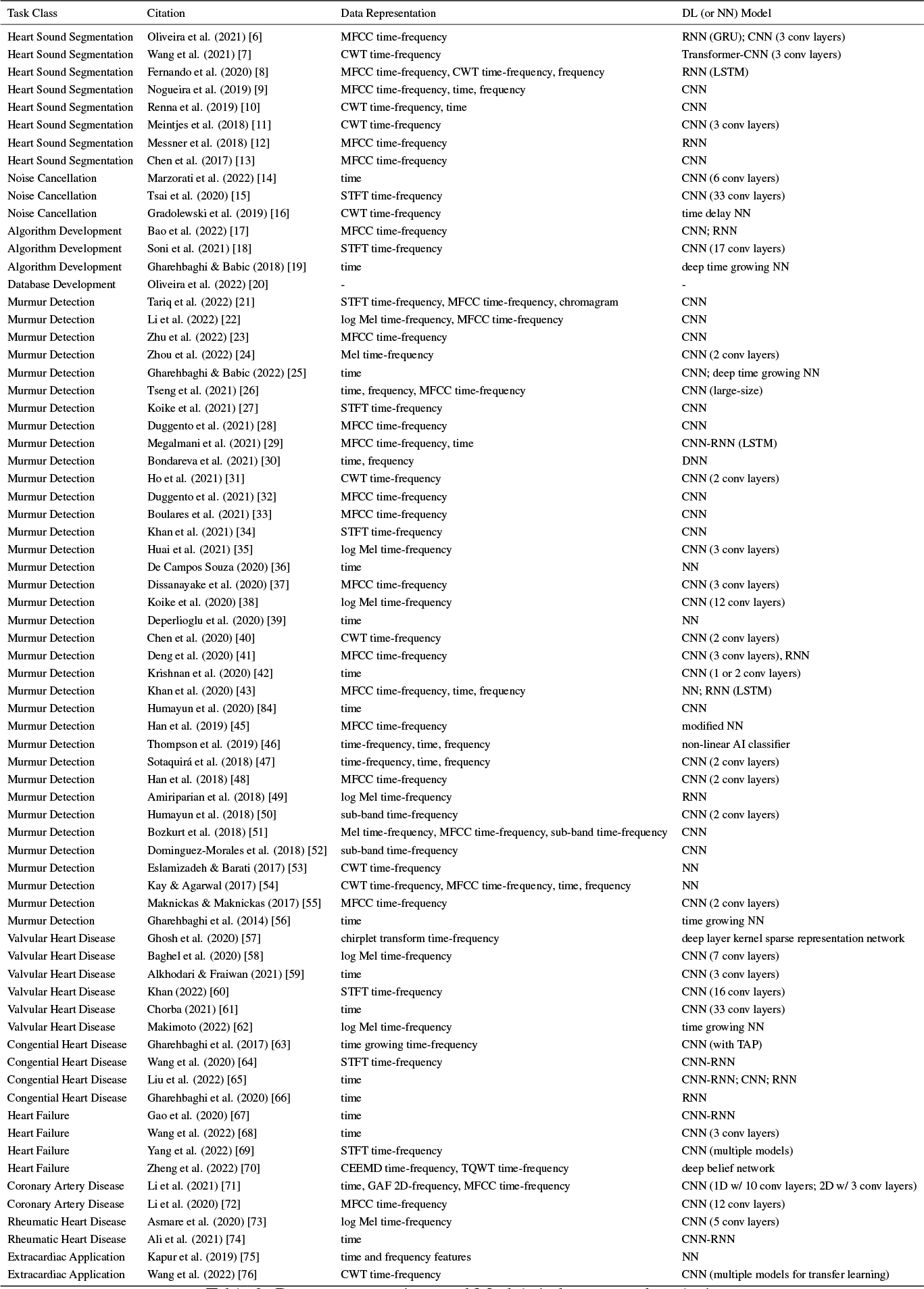
Data representations and Models in heart sound analysis.

### 3.3 Applications

As shown in Figure 1, except for a few studies focusing on method development such as segmentation, noise cancellation, algorithm development, and database development, most applications of DL heart sound analysis are related to the clinical field, and we will discuss the details below.

#### 3.3.1 Cardiac murmurs detection

Detecting cardiac murmurs is a fundamental but crucial task in heart sound analysis. Cardiac murmurs are abnormal sounds produced during the cardiac cycle and can indicate underlying heart conditions. As previously indicated, identifying cardiac murmurs through auscultation demands a combination of expertise and experience, and can lead to variability across different specialists. Since detecting cardiac murmurs is a relatively more straightforward task compared to identifying specific diseases, there has been a notable accumulation of high-quality annotated data in recent years, which has been made available to the public [20, 94]. The abundance of accessible data has led to a surge in DL models research on cardiac murmurs detection [21, 22, 23, 24, 25, 26, 27, 28, 29, 30, 31, 32, 33, 34, 35, 36, 37, 38, 39, 40, 41, 42, 43, 44, 45, 46, 47, 48, 49, 50, 51, 52, 53, 54, 55, 56]. Although these models can only discern the presence of cardiac murmurs and cannot provide definitive diagnoses, they still play a crucial role in community-based disease screening. This facilitates referring individuals with potential cardiac conditions to specialists for a precise diagnosis. Such an approach holds immense value, especially in underprivileged regions with limited medical resources.

#### 3.3.2 Valvular heart disease

Valvular heart disease (VHD) is a prevalent condition associated with high mortality rates worldwide [95]. Early screening and follow-up are crucial for managing VHD as most patients remain asymptomatic until the advanced stage, resulting in poor prognosis without timely intervention. While echocardiography is the current gold standard for VHD diagnosis [96, 97], its cost and requirement for specialized personnel make it impractical for community screening and self-monitoring.

Cardiac auscultation is a simple and cost-effective diagnostic tool for VHD. However, relying solely on auscultation for diagnosis results in low accuracy due to human errors and environmental disturbances [98]. DL techniques have demonstrated superior recognition capabilities compared to humans. Using the Khan dataset [80] consisting of 1,000 audio clips of normal heart sounds and four VHDs: aortic stenosis (AS), mitral stenosis (MS), mitral regurgitation (MR), and mitral valve prolapse (MVP), several DL algorithms have been developed to extract heart sound features and train models for VHD diagnosis. Yaseen et al. employed Mel-frequency Cepstral Coefficients (MFCCs) combined with Discrete Wavelet Transform features as inputs to Deep Neural Network (DNN) classifiers, achieving an accuracy of 92.1% [80]. Ghosh et al. extracted features from the time-frequency matrix of the heart sound recordings and input them into a Deep Layer Kernel Sparse Representation Network classifier, resulting in a 99.24% accuracy [57]. Abbas et al. developed a novel attention-based transformer architecture that combines DL and vision transformer. They utilized the continuous wavelet transform-based spectrogram strategy to extract representative features and built an attention-based model on a convolutional vision transformer, yielding an overall average accuracy of 100%, sensitivity of 99.0%, Specificity of 99.5%, and F1-score of 98.0% [99].

Recently, researchers have worked on constructing automatic models that do not require signal pre-processing or feature engineering. Also based on the Khan dataset [80], Baghel et al. employed a CNN with data augmentation and a Gaussian filter for noise removal, achieving an accuracy of 98.6% [58]. Alkhodari et al. utilized a CNN-RNN model for direct classification using heart sound recordings with one-dimensional wavelet smoothing, resulting in a 99.32% accuracy [59]. Khan et al. developed a novel Cardi-Net architecture based on a CNN structure to extract discriminative PCG features from the power spectrogram for VHD identification, achieving an accuracy of 98.88% [60]. In a large-scale study, Chorba et al. trained a CNN model on over 34 hours of heart sound recordings from 5,318 patients to detect VHD-related murmurs, yielding promising performance [61]. Makimoto and Shiraga et al. further improved the interpretability and usability of DL models. They developed a lightweight CNN model to detect severe AS using 1,668 heart sound recordings at three auscultation locations from 556 patients. Based on this model, a smartphone application was established, achieving a 95.7% accuracy and 0.93 F1 score. Additionally, they employed Gradient-based Class Activation Maps to identify the specific heart sound features that the DL model focused on when distinguishing the severity of AS [62].

#### 3.3.3 Congenital heart disease

Congenital heart disease (CHD) is a prevalent cardiovascular disease in children, affecting approximately 0.8-1% global population [100]. The most common type is left-to-right shunt CHD, including atrial septal defects (ASD), ventricular septal defects (VSD), and patent ductus arteriosus (PDA). This condition can cause chronic volume overload, resulting in heart failure and pulmonary hypertension [101, 102]. Imaging techniques, including echocardiography, magnetic resonance imaging, and computerized tomography, are crucial for CHD evaluation [103]. However, their limited availability and high costs pose challenges, particularly in underdeveloped regions. The delay in diagnosis can lead to irreversible complications and even death [104].

Auscultation plays a vital role in screening and diagnosing CHD as these patients often present with heart murmurs caused by abnormal blood flow through malformed heart structures[105]. However, the accuracy of this method heavily relies on the physicians’ experience, and not all heart murmurs can be accurately identified [106]. To enhance the diagnostic efficiency of heart sound, DL algorithms have been increasingly employed. Wang et al. developed a temporal attentive pooling-convolutional recurrent neural network model for VSD detection using heart sound recordings from 51 patients with VSD and 25 healthy individuals, with a sensitivity of 96.0% and a specificity of 96.7%. Notably, when analyzing heart sounds in the second aortic and tricuspid areas, the sensitivity and specificity of this model reached 100% [64]. Huang et al. converted heart sound recordings from 184 participants, including 46 with VSDs, 50 with ASDs, and 88 with a normal heart structure, into bispectrum signals. These signals were then utilized to train an advanced optical coherence tomography network model for heart sound classification. Remarkably, this model outperformed experienced cardiologists in detecting VSD and ASD with an accuracy of 93.4% and 85.3%, respectively [107]. Liu et al. developed a residual convolution recurrent neural network model to detect ASD, VSD, PDA, and combined CHD using 884 heart sound recordings from children with left-to-right shunt CHD, with accuracy values ranging from 94.0% to 99.4% [65].

In clinical practice, distinguishing between VSD and bicuspid/tricuspid regurgitation through auscultation can be challenging, as both conditions manifest as systolic murmurs. Gharehbaghi et al. tackled this issue by training a Time Growing Neural Network (TGNN) model to differentiate VSD from valvular regurgitation and healthy subjects using heart sound recordings from 90 individuals, achieving an accuracy of 86.7% [63]. Furthermore, innocent murmurs are present in approximately 50% of children, leading to a significant number of unnecessary referrals to pediatric cardiologists [108]. To address the issue, Gharehbaghi et al. developed a TGNN model capable of distinguishing ASD and VSD from valvular regurgitation and innocent murmur using heart sound recordings from 115 children, resuting in an accuracy of 91.6% [66].

#### 3.3.4 Heart failure

Heart failure (HF) is a global epidemic with high mortality, affecting over 26 million individuals worldwide, and its prevalence continues to rise due to an aging population [109]. Early detection and timely treatment of HF are crucial for long-term prognosis, as the progression of HF can lead to irreversible myocardial remodeling and functional impairment [110]. Current guidelines outline specific conditions for the diagnosis of HF, including typical symptoms and signs, reduced or preserved LVEF, elevated brain natriuretic peptide levels, and the presence of structural heart disease and diastolic dysfunction [111, 112],. However, the symptoms or signs may be non-specific at the early stages of HF [111, 112], and echocardiography and blood biomarker tests are unsuitable for screening purposes.

Heart sounds, as a physiological signal generated by myocardial contraction, can provide direct insights into the mechanical dysfunctions of the heart [113]. However, the heart sounds specific to HF, such as gallop rhythm, usually become apparent at the later stages of HF and require sufficient expertise to identify. With DL algorithms, Gao et al. first proposed an HF screening framework based on a gated recurrent unit (GRU) model, distinguishing between the normal subjects, HF with preserved ejection fraction (HFpEF), and HF with reduced ejection fraction (HFrEF) using heart sounds, with an average accuracy of 98.82% [67]. Wang et al. employed CNN and RNN to build a heart sound diagnostic model that accurately differentiated between normal individuals, HFpEF, and HFrEF, achieving an accuracy of 97.64% [68]. Yang et al. developed a CNN model to diagnose left ventricular diastolic dysfunction using heart sounds. They utilized data augmentation techniques with deep convolutional generative adversarial networks to enhance the model performance, resulting in an accuracy of 98.7% [69].

Once HF is diagnosed, accurately classifying the stages of HF is critical for guiding clinical practice. The AHA/ACC guidelines define four stages of HF (stages A, B, C, and D), ranging from developing HF without symptoms to advanced HF [112]. Zheng et al. utilized heart sound recordings from 275 subjects and employed a deep belief network model that incorporated multi-scale (original signal, sub-sequences, and sub-band signals) and multi-domain (time-domain, frequency-domain, and nonlinear) features. Their approach achieved 74.3% accuracy in automatically HF staging [70].

#### 3.3.5 Coronary artery disease

Coronary artery disease (CAD) is a major cause of mortality and morbidity worldwide and substantially burdens the medical system [114]. While coronary angiography is considered the gold standard for CAD diagnosis, its invasive nature and requirement for specialized catheterization laboratories restrict it availability. Electrocardiogram is another commonly used diagnostic tool, but it has limitations in terms of sensitivity, particularly in stable and asymptomatic patients, and its accuracy highly depends on the expertise of the interpreting physicians [115]. Consequently, there may be a considerable number of undiagnosed CAD cases in underdeveloped regions.

Previous studies have shown that turbulence in stenosed coronary arteries can produce faint high-frequency murmurs [116, 117, 118]. However, these faint murmurs are often not discernible during auscultation, and recognizable changes in heart sounds typically occur only after the development of severe structural complications, such as papillary muscle dysfunction, septal perforation, or ventricular dilatation [1]. Given that machine recording can capture faint murmurs, a DL-based diagnostic model shows promise for CAD detection. In 2020, Li et al. developed a CAD detection model using heart sounds. They extracted 110 multi-domain features and MFCCs from the heart sound recordings of 175 subjects. The fusion framework, combining selected multi-domain and DL features, served as input for a CNN classifier, achieving an accuracy of 90.43% [72]. In 2021, Li et al. further improved their approach by developing a multi-input CNN framework that integrated time, frequency, and time-frequency domain deep features from simultaneous electrocardiogram and PCG signals of 195 subjects for CAD detection. The model, which combined multi-domain deep features from the two modalities, showed high performance in CAD identification, with an accuracy of 96.51% [71]. To address the challenge of limited sample size for training CNN models, Pathak et al. explored transfer learning for CAD detection using heart sounds. They employed a CNN pre-trained on the ImageNet database, consisting of 1 million training images, and transferred its feature representation for CAD detection. Multiple kernel learning was then used to fuse the embeddings of the CNN with handcrafted features, including the heat map of Synchrosqueezing Transform, time-varying Shannon and Renyi Entropy in subbands of Synchrosqueezing Transform. Despite having only 40 CAD and 40 normal subjects’ heart sound data, their diagnostic model achieved an accuracy of 89.25% [119].

#### 3.3.6 Rheumatic heart disease

Rheumatic heart disease (RHD) remains a significant public health issue in developing countries, impacting a minimum of 33 million individuals and contributing to at least 345,000 deaths annually [120]. RHD is caused by an abnormal immune response to beta-hemolytic streptococcal pharyngitis infection and primarily affects the mitral valve. Typically, echocardiography is used to diagnose RHD by evaluating valve morphology and severity of valve dysfunction [121]. However, given the high prevalence of RHD in underdeveloped regions, there is an urgent need to develop a cost-effective screening method for RHD.

The RHD-related damage on the valves disrupts the normal blood flow in the heart chambers and causes murmurs, which presents a possibility of creating a DL-based model for RHD detection using heart sounds. In 2020, Asmare et al. collected 33453 heart sound clips from 124 RHD patients and 46 healthy individuals. They trained a CNN model using the Mel Spectro-temporal representation of un-segmented PCG, achieving an overall accuracy of 96.1% with 94.0% sensitivity and 98.1% specificity [73].

Compared to intervention after RHD has developed, early detection of subclinical RHD in susceptible populations and providing penicillin for prophylaxis may be a more cost-effective strategy for individuals and healthcare systems [122]. Based on this principle, Ali et al. proposed a study plan to recruit 1700 children (5–15 years) from underprivileged schools in Pakistan and collect clinical data, including heart sound recordings and echocardiograms. They aimed to train a DNN to automatically identify patients with subclinical RHD and definite RHD [74]. This study is currently ongoing, and we look forward to its results.

#### 3.3.7 Extracardiac applications

Continuous blood pressure (BP) measurements are essential for managing critically ill patients and those undergoing surgery. Invasive intra-arterial cannulation is the gold standard for continuous BP measurement. However, it often causes arterial complications and thrombosis. On the other hand, the cuff BP measurement, the most common non-invasive method, can provide only indirect estimates of systolic and diastolic BP using proprietary formulas, and it does not allow for continuous readings [123]. Heart sounds have shown a close relationship with BP. Previous studies have established a positive correlation between the frequency and amplitude of the second heart sound and BP [124, 125]. This relationship can be explained by the mechanical vibrations caused by arterial wall elasticity and blood column inertia [126]. Additionally, the amplitude of the first heart sound has been linked to cardiac contractility [126]. These findings provide a physiological basis for estimating BP using heart sounds with DL techniques. In 2019, Kapur et al. trained an artificial neural network model to estimate BP using 737 heart sound recordings from 25 children undergoing continuous BP monitoring via radial artery intra-arterial catheters. The DL model successfully estimated BP, exhibiting a significant correlation with the readings obtained from intra-arterial catheters (R2 = 0.928 and 0.868 for systolic and diastolic BP, respectively) [75].

Pulmonary hypertension (PH) is a chronic and progressive disease characterized by dyspnea, right heart failure, and a high mortality risk [127]. The gold standard for diagnosing PH is right heart catheterization, which defines PH as a resting mean pulmonary artery pressure (PAP) of 20 mmHg or higher. However, cardiac catheterization is invasive and thus unsuitable for routine examinations. As an alternative, echocardiography is recommended for estimating PAP, calculated from the maximum peak tricuspid regurgitation velocity using the Bernoulli equation. Nonetheless, echocardiography is operator-dependent and requires optimal acoustic windows and flow tracings to measure PAP accurately, resulting in a delay of up to 2 years between the symptom onset and PH diagnosis [128]. As PAP increases, specific changes occur in heart sounds, including tricuspid regurgitant murmurs, an augmented second heart sound in the pulmonic area, and a third heart sound gallop. Wang et al. utilized the Khan dataset [80] and supplemented it with their own heart sound recordings of PH. They translated one-dimensional heart sound signals into three-dimensional spectrograms using CWT. They employed ten transfer learning networks to diagnose PH and VHDs, and compared their performance. Their findings revealed that four transfer learning networks (ResNet101, DenseNet201, DarkNet19, and GoogleNet) outperformed other models with an accuracy of 98% in detecting PH and 4 VHDs [76].

## 4 Discussion

DL has immense potential in analyzing heart sounds, enabling precise and automated diagnosis of heart conditions. However, this field also presents several challenges and opportunities for further development.

### 4.1 Data limitation

The limited data availability is one challenge in using DL for heart sound analysis. Heart sound data collection and annotation is complex and time-consuming, requiring specialized equipment and trained clinicians. Consequently, the quantity of labeled heart sound data is considerably smaller than other medical data types, such as medical images or electronic health records (see Table 2). However, DL models thrive on large-scale datasets to learn and generalize effectively. Insufficient data may result in overfitting, where the model memorizes the available examples instead of learning meaningful features, leading to poor generalization of unseen data. Leveraging transfer learning techniques can be beneficial when faced with limited heart sound data. Pre-training DL models on large-scale datasets from related domains, such as general audio data or medical imaging, can help initialize the model with useful features, enabling it to learn from limited labeled heart sound data more effectively [76, 119].

The quality of heart sound recordings is another challenge for DL model construction. The acquisition of heart sounds in real-world practice is vulnerable to interference from environmental noise, which may obscure faint murmurs and degrade the quality of recordings [58]. Additionally, the positioning of the stethoscope during data collection can significantly influence the characteristics of recorded heart sounds [1], subsequently affecting the performance of DL models trained on such data. Therefore, it is crucial to investigate and establish standardized and rigorous protocols for heart sound collection to ensure consistent and reliable results.

### 4.2 Pre-processing disadvantage

As previously mentioned, heart sounds are typically pre-processed to extract their frequency information and represent it in a time-frequency format, such as a spectrogram or Mel-spectrogram. This representation is then fed into a DL model, which can identify patterns and features associated with various heart conditions. Although this pre-processing step might make the model converge quicker and incorporate prior knowledge from engineering or human hearing principles, it also risks filtering out valuable information that could benefit the model’s prediction. Numerous studies mentioned above have delved into various signal-processing techniques. However, a comprehensive investigation into the impact of signal processing on the model’s overall performance is still lacking. It is essential to conduct further research and exploration to understand the trade-off between prior knowledge and detailed information in heart sound analysis when employing DL techniques.

### 4.3 Interpretability shortage

Like with the application of DL in other medical fields, the interpretability of DL in heart sound analysis is limited. As DL models are designed to handle the complexities and nuances of large datasets, they are often too complex to comprehend or explain fully. Therefore, it is difficult to determine why a given model may be producing a certain result or why it may be missing specific nuances in the data. Furthermore, DL models are prone to overfitting, leading to results specific to a given dataset but not generalizable to other datasets. As such, there is an inherent need for caution when using DL for heart sound analysis. Despite these limitations, DL models have been successfully applied to heart sound analysis, yielding promising results. As mentioned above, they have shown superiority over traditional signal processing techniques in detecting complex cardiovascular events using heart sound data, contributing to advancements in the field and holding potential for improving diagnostic accuracy and automation in cardiac healthcare.

### 4.4 Future perspectives

#### Smartphone applications

The increased computing power of smartphones has made them well-suited for deploying DL-based heart sound diagnostic models. Smartphones come equipped with built-in microphones that can capture heart sounds of sufficient quality. In a study conducted by Luo et al. using their smartphone application, over 80% of users were able to obtain good-quality heart sound recordings, with success rates independent of age, gender, body mass index, and smartphone versions [129]. As mentioned above, Makimoto et al. have developed a smartphone application using a lightweight CNN model to detect severe AS [62]. Smartphone-based diagnostic models offer a convenient way for patients to monitor their health status at home, enabling the early detection of potential diseases and ensuring prompt access to treatment. Such applications also democratize healthcare by providing individuals, regardless of their geographic location or socioeconomic status, with the same opportunity to perform disease screenings using their smartphones.

#### Wearable devices

Flexible heart sound sensors, such as fabric material sensors, offer greater convenience and comfort than traditional sensors, allowing for long-term wear and heart sound recording [130]. These continuous monitoring data can be processed using DL models to detect anomalies and predict adverse events in real time. For instance, patients with HF can take advantage of wearable heart sound collection devices, which facilitate continuous monitoring of their cardiac function. This enables the early detection of acute exacerbations of HF, thereby prompting timely medical intervention. To the best of our knowledge, there are no DL models specifically designed to process long-term heart sound data. However, as wearable devices continue to advance, the accumulation of relevant data will propel the development of these specialized DL models.

#### Multi-modalities

The integration of different modalities in DL models can uncover hidden patterns and dependencies that might not be apparent when analyzing each modality individually. Li et al. proposed a multi-modal machine learning approach that combines simultaneous electrocardiogram and PCG signals to predict cardiovascular diseases [131]. Their study demonstrated that the performance of the multi-modal method surpassed that of single models based solely on electrocardiogram or PCG. However, there is no research on the multi-modal analysis of PCG specifically using DL networks. The combination of multi-modal DL-based heart sound diagnostic models holds promise to further enhance diagnostic accuracy.

## 5 Conclusion

Cardiac auscultation is a fundamental and essential skill for clinicians, but it requires extensive training and experience to identify and diagnose heart conditions accurately. Nowadays, heart sounds can be easily recorded and analyzed using computers. By combining the traditional signal processing approaches and DL techniques, researchers have made significant progress in detecting a wide range of cardiovascular diseases using heart sounds. While some promising results have been achieved using DL models for diagnosing heart conditions based on PCG data, further research is needed to validate the accuracy and generalizability of these models.

## Data Availability

The data availability is based on each literature discussed in the review.

## Funding

This work was supported by the National Natural Science Foundation of China (No. 62102008) and Peking University People’s Hospital Scientific Research Development Funds (RDJP2022-39).

## Abbreviations

DL: deep learning
PCG: phonocardiogram
AS: aortic stenosis
MR: mitral regurgitation
MS: mitral stenosis
MVP: mitral valve prolapse
STFT: Short-Time Fourier-Transform
MFCCs: Mel-frequency Cepstral Coefficients
CEEMD: Complementary Ensemble Empirical Mode Decomposition
TQWT: Tunable-Q Wavelet Transform
CWT: Continuous Wavelet Transform
CNNs: convolutional neural networks
RNNs: recurrent neural networks
NN: neural network
ML: machine learning
VHD: Valvular heart disease
DNN: Deep Neural Network
CHD: Congenital heart disease
ASD: atrial septal defects
VSD: ventricular septal defects
PDA: patent ductus arteriosus
TGNN: Time Growing Neural Network
LVEF: left ventricular ejection fraction
GRU: gated recurrent unit
HFpEF: Heart failure with preserved ejection fraction
HFrEF: Heart failure with reduced ejection fraction
LVDD: left ventricular diastolic dysfunction
CAD: Coronary artery disease
RHD: Rheumatic heart disease
BP: blood pressure
PH: pulmonary hypertension
PAP: pulmonary artery pressure

## Notes

### Competing Interest Statement

The authors have declared no competing interest.

### Funding Statement

This study was funded by the National Natural Science Foundation of China (No. 62102008) and Peking University People's Hospital Scientific Research Development Funds (RDJP2022-39).

## References

[1] Zipes DP. Braunwald’s Heart Disease: A Textbook of Cardiovascular Medicine;5(2):63–3.

[2] Etchells E, Bell C, Robb K. Does This Patient Have an Abnormal Systolic Murmur?;277(7):564–71.

[3] Mangione S, Nieman LZ. Cardiac Auscultatory Skills of Internal Medicine and Family Practice Trainees: A Comparison of Diagnostic Proficiency;278(9):717–22.

[4] Chen W, Sun Q, Chen X, Xie G, Wu H, Xu C. Deep Learning Methods for Heart Sounds Classification: A Systematic Review;23(6):667.

[5] Li S, Li F, Tang S, Xiong W. A Review of Computer-Aided Heart Sound Detection Techniques;2020:1–10.

[6] Oliveira J, Nogueira D, Renna F, Ferreira C, Jorge AM, Coimbra M. Do We Really Need a Segmentation Step in Heart Sound Classification Algorithms? In: 2021 43rd Annual International Conference of the IEEE Engineering in Medicine & Biology Society (EMBC). IEEE;. p. 286–9.

[7] Wang X, Liu C, Li Y, Cheng X, Li J, Clifford GD. Temporal-framing adaptive network for heart sound segmentation without prior knowledge of state duration. IEEE Transactions on Biomedical Engineering. 2020;68(2):650–63.

[8] Fernando T, Ghaemmaghami H, Denman S, Sridharan S, Hussain N, Fookes C. Heart sound segmentation using bidirectional LSTMs with attention. IEEE journal of biomedical and health informatics. 2019;24(6):1601–9.

[9] Nogueira DM, Ferreira CA, Gomes EF, Jorge AM. Classifying Heart Sounds Using Images of Motifs, MFCC and Temporal Features;43(6):168.

[10] Renna F, Oliveira J, Coimbra MT. Deep Convolutional Neural Networks for Heart Sound Segmentation;23(6):2435–45.

[11] Meintjes A, Lowe A, Legget M. Fundamental Heart Sound Classification Using the Continuous Wavelet Transform and Convolutional Neural Networks. In: 2018 40th Annual International Conference of the IEEE Engineering in Medicine and Biology Society (EMBC). IEEE;. p. 409–12.

[12] Messner E, Zohrer M, Pernkopf F. Heart Sound Segmentation—An Event Detection Approach Using Deep Recurrent Neural Networks;65(9):1964–74.

[13] Chen TE, Yang SI, Ho LT, Tsai KH, Chen YH, Chang YF, et al. S1 and S2 heart sound recognition using deep neural networks. IEEE Transactions on Biomedical Engineering. 2016;64(2):372–80.

[14] Marzorati D, Dorizza A, Bovio D, Salito C, Mainardi L, Cerveri P. Hybrid Convolutional Networks for End-to-End Event Detection in Concurrent PPG and PCG Signals Affected by Motion Artifacts;69(8):2512–23.

[15] Tsai KH, Wang WC, Cheng CH, Tsai CY, Wang JK, Lin TH, et al. Blind Monaural Source Separation on Heart and Lung Sounds Based on Periodic-Coded Deep Autoencoder;24(11):3203–14.

[16] Gradolewski D, Magenes G, Johansson S, Kulesza W. A Wavelet Transform-Based Neural Network Denoising Algorithm for Mobile Phonocardiography;19(4):957.

[17] Bao X, Xu Y, Kamavuako EN. The Effect of Signal Duration on the Classification of Heart Sounds: A Deep Learning Approach;22(6):2261.

[18] Soni PN, Shi S, Sriram PR, Ng AY, Rajpurkar P. Contrastive Learning of Heart and Lung Sounds for Label-Efficient Diagnosis;3(1):100400.

[19] Gharehbaghi A, Babic A. Structural risk evaluation of a deep neural network and a Markov model in extracting medical information from phonocardiography. In: Data, Informatics and Technology: An Inspiration for Improved Healthcare. IOS Press; 2018. p. 157–60.

[20] Oliveira J, Renna F, Costa PD, Nogueira M, Oliveira C, Ferreira C, et al. The CirCor DigiScope dataset: from murmur detection to murmur classification. IEEE journal of biomedical and health informatics. 2021;26(6):2524–35.

[21] Tariq Z, Shah SK, Lee Y. Feature-based fusion using CNN for lung and heart sound classification. Sensors. 2022;22(4):1521.

[22] Li Z, Chang Y, Schuller BW. CNN-Based Heart Sound Classification with an Imbalance-Compensating Weighted Loss Function. In: 2022 44th Annual International Conference of the IEEE Engineering in Medicine & Biology Society (EMBC). IEEE; 2022. p. 4934–7.

[23] Zhu L, Qian K, Wang Z, Hu B, Yamamoto Y, Schuller BW. Heart Sound Classification Based on Residual Shrinkage Networks. In: 2022 44th Annual International Conference of the IEEE Engineering in Medicine & Biology Society (EMBC). IEEE;. p. 4469–72.

[24] Zhou G, Chen Y, Chien C. On the Analysis of Data Augmentation Methods for Spectral Imaged Based Heart Sound Classification Using Convolutional Neural Networks;22(1):226.

[25] Gharehbaghi A, Babic A. Deep Time Growing Neural Network vs Convolutional Neural Network for Intelligent Phonocardiography. In: Mantas J, Gallos P, Zoulias E, Hasman A, Househ MS, Diomidous M, et al., editors. Studies in Health Technology and Informatics. IOS Press;. .

[26] Tseng KK, Wang C, Huang YF, Chen GR, Yung KL, Ip WH. Cross-domain transfer learning for pcg diagnosis algorithm. Biosensors. 2021;11(4):127.

[27] Koike T, Qian K, Schuller BW, Yamamoto Y. Transferring Cross-Corpus Knowledge: An Investigation on Data Augmentation for Heart Sound Classification. In: 2021 43rd Annual International Conference of the IEEE Engineering in Medicine & Biology Society (EMBC). IEEE;. p. 1976–9.

[28] Duggento A, Conti A, Guerrisi M, Toschi N. Classification of Real-World Pathological Phonocardiograms through Multi-Instance Learning. In: 2021 43rd Annual International Conference of the IEEE Engineering in Medicine & Biology Society (EMBC). IEEE;. p. 771–4.

[29] Megalmani DR, G SB, Rao M V A, Jeevannavar SS, Ghosh PK. Unsegmented Heart Sound Classification Using Hybrid CNN-LSTM Neural Networks. In: 2021 43rd Annual International Conference of the IEEE Engineering in Medicine & Biology Society (EMBC). IEEE;. p. 713–7.

[30] Bondareva E, Han J, Bradlow W, Mascolo C. Segmentation-Free Heart Pathology Detection Using Deep Learning. In: 2021 43rd Annual International Conference of the IEEE Engineering in Medicine & Biology Society (EMBC). IEEE;. p. 669–72.

[31] Ho WH, Huang TH, Yang PY, Chou JH, Qu JY, Chang PC, et al. Robust Optimization of Convolutional Neural Networks with a Uniform Experiment Design Method: A Case of Phonocardiogram Testing in Patients with Heart Diseases;22(S5):92.

[32] Duggento A, Conti A, Guerrisi M, Toschi N. A Novel Multi-Branch Architecture for State of the Art Robust Detection of Pathological Phonocardiograms;379(2212):20200264.

[33] Boulares M, Alotaibi R, AlMansour A, Barnawi A. Cardiovascular Disease Recognition Based on Heartbeat Segmentation and Selection Process;18(20):10952.

[34] Khan KN, Khan FA, Abid A, Olmez T, Dokur Z, Khandakar A, et al. Deep Learning Based Classification of Unsegmented Phonocardiogram Spectrograms Leveraging Transfer Learning;42(9):095003.

[35] Huai X, Kitada S, Choi D, Siriaraya P, Kuwahara N, Ashihara T. Heart Sound Recognition Technology Based on Convolutional Neural Network;46(3):320–32.

[36] de Campos Souza PV, Lughofer E. Identification of Heart Sounds with an Interpretable Evolving Fuzzy Neural Network. Sensors. 2020;20(22). Available from: https://www.mdpi.com/1424-8220/20/22/6477.

[37] Dissanayake T, Fernando T, Denman S, Sridharan S, Ghaemmaghami H, Fookes C. A robust interpretable deep learning classifier for heart anomaly detection without segmentation. IEEE Journal of Biomedical and Health Informatics. 2020;25(6):2162–71.

[38] Koike T, Qian K, Kong Q, Plumbley MD, Schuller BW, Yamamoto Y. Audio for Audio Is Better? An Investigation on Transfer Learning Models for Heart Sound Classification. In: 2020 42nd Annual International Conference of the IEEE Engineering in Medicine & Biology Society (EMBC). IEEE;. p. 74–7.

[39] Deperlioglu O, Kose U, Gupta D, Khanna A, Sangaiah AK. Diagnosis of Heart Diseases by a Secure Internet of Health Things System Based on Autoencoder Deep Neural Network;162:31–50.

[40] Chen Y, Wei S, Zhang Y. Classification of Heart Sounds Based on the Combination of the Modified Frequency Wavelet Transform and Convolutional Neural Network;58(9):2039–47.

[41] Deng M, Meng T, Cao J, Wang S, Zhang J, Fan H. Heart Sound Classification Based on Improved MFCC Features and Convolutional Recurrent Neural Networks;130:22–32.

[42] Krishnan PT, Balasubramanian P, Umapathy S. Automated Heart Sound Classification System from Unsegmented Phonocardiogram (PCG) Using Deep Neural Network;43(2):505–15.

[43] Khan FA, Abid A, Khan MS. Automatic Heart Sound Classification from Segmented/Unsegmented Phonocardiogram Signals Using Time and Frequency Features;41(5):055006.

[44] Humayun AI, Ghaffarzadegan S, Ansari MI, Feng Z, Hasan T. Towards domain invariant heart sound abnormality detection using learnable filterbanks. IEEE journal of biomedical and health informatics. 2020;24(8):2189–98.

[45] Han W, Xie S, Yang Z, Zhou S, Huang H. Heart Sound Classification Using the SNMFNet Classifier;40(10):105003.

[46] Thompson WR, Reinisch AJ, Unterberger MJ, Schriefl AJ. Artificial Intelligence-Assisted Auscultation of Heart Murmurs: Validation by Virtual Clinical Trial;40(3):623–9.

[47] Sotaquirá M, Alvear D, Mondragón M. Phonocardiogram Classification Using Deep Neural Networks and Weighted Probability Comparisons;42(7):510–7.

[48] Han W, Yang Z, Lu J, Xie S. Supervised Threshold-Based Heart Sound Classification Algorithm;39(11):115011.

[49] Amiriparian S, Schmitt M, Cummins N, Qian K, Dong F, Schuller B. Deep Unsupervised Representation Learning for Abnormal Heart Sound Classification. In: 2018 40th Annual International Conference of the IEEE Engineering in Medicine and Biology Society (EMBC). IEEE;. p. 4776–9.

[50] Humayun AI, Ghaffarzadegan S, Feng Z, Hasan T. Learning Front-end Filter-bank Parameters Using Convolutional Neural Networks for Abnormal Heart Sound Detection. In: 2018 40th Annual International Conference of the IEEE Engineering in Medicine and Biology Society (EMBC);. p. 1408–11.

[51] Bozkurt B, Germanakis I, Stylianou Y. A Study of Time-Frequency Features for CNN-based Automatic Heart Sound Classification for Pathology Detection;100:132–43.

[52] Dominguez-Morales JP, Jimenez-Fernandez AF, Dominguez-Morales MJ, Jimenez-Moreno G. Deep neural networks for the recognition and classification of heart murmurs using neuromorphic auditory sensors. IEEE transactions on biomedical circuits and systems. 2017;12(1):24–34.

[53] Eslamizadeh G, Barati R. Heart Murmur Detection Based on Wavelet Transformation and a Synergy between Artificial Neural Network and Modified Neighbor Annealing Methods;78:23–40.

[54] Kay E, Agarwal A. DropConnected Neural Networks Trained on Time-Frequency and Inter-Beat Features for Classifying Heart Sounds;38(8):1645–57.

[55] Maknickas V, Maknickas A. Recognition of Normal–Abnormal Phonocardiographic Signals Using Deep Convolutional Neural Networks and Mel-Frequency Spectral Coefficients;38(8):1671–84.

[56] Gharehbaghi A, Dutoit T, Ask P, Sörnmo L. Detection of Systolic Ejection Click Using Time Growing Neural Network;36(4):477–83.

[57] Ghosh SK, Ponnalagu RN, Tripathy RK, Acharya UR. Deep Layer Kernel Sparse Representation Network for the Detection of Heart Valve Ailments from the Time-Frequency Representation of PCG Recordings;2020:1–16.

[58] Baghel N, Dutta MK, Burget R. Automatic Diagnosis of Multiple Cardiac Diseases from PCG Signals Using Convolutional Neural Network;197:105750.

[59] Alkhodari M, Fraiwan L. Convolutional and Recurrent Neural Networks for the Detection of Valvular Heart Diseases in Phonocardiogram Recordings;200:105940.

[60] Khan JS, Kaushik M, Chaurasia A, Dutta MK, Burget R. Cardi-Net: A Deep Neural Network for Classification of Cardiac Disease Using Phonocardiogram Signal;219:106727.

[61] Chorba JS, Shapiro AM, L. L, Maidens J, Prince J, Pham S, et al. Deep Learning Algorithm for Automated Cardiac Murmur Detection via a Digital Stethoscope Platform;10(9):e019905.

[62] Makimoto H, Shiraga T, Kohlmann B, Magnisali CE, Gerguri S, Motoyama N, et al. Efficient Screening for Severe Aortic Valve Stenosis Using Understandable Artificial Intelligence: A Prospective Diagnostic Accuracy Study;3(2):141–52.

[63] Gharehbaghi A, Sepehri AA, Linden M, Babic A. Intelligent Phonocardiography for Screening Ventricular Septal Defect Using Time Growing Neural Network.;. p. 108–11.

[64] Wang JK, Chang YF, Tsai KH, Wang WC, Tsai CY, Cheng CH, et al. Automatic Recognition of Murmurs of Ventricular Septal Defect Using Convolutional Recurrent Neural Networks with Temporal Attentive Pooling;10(1):21797.

[65] Liu J, Wang H, Yang Z, Quan J, Liu L, Tian J. Deep Learning-Based Computer-Aided Heart Sound Analysis in Children with Left-to-Right Shunt Congenital Heart Disease;348:58–64.

[66] Gharehbaghi A, Sepehri AA, Babic A. Distinguishing Septal Heart Defects from the Valvular Regurgitation Using Intelligent Phonocardiography.

[67] Gao S, Zheng Y, Guo X. Gated Recurrent Unit-Based Heart Sound Analysis for Heart Failure Screening;19(1):3.

[68] Wang H, Guo X, Zheng Y, Yang Y. An Automatic Approach for Heart Failure Typing Based on Heart Sounds and Convolutional Recurrent Neural Networks;45(2):475–85.

[69] Yang Y, Guo XM, Wang H, Zheng YN. Deep Learning-Based Heart Sound Analysis for Left Ventricular Diastolic Dysfunction Diagnosis;11(12):2349.

[70] Zheng Y, Guo X, Wang Y, Qin J, Lv F. A Multi-Scale and Multi-Domain Heart Sound Feature-Based Machine Learning Model for ACC/AHA Heart Failure Stage Classification;43(6):065002.

[71] Li H, Wang X, Liu C, Li P, Jiao Y. Integrating Multi-Domain Deep Features of Electrocardiogram and Phonocardiogram for Coronary Artery Disease Detection;138:104914.

[72] Li H, Wang X, Liu C, Zeng Q, Zheng Y, Chu X, et al. A Fusion Framework Based on Multi-Domain Features and Deep Learning Features of Phonocardiogram for Coronary Artery Disease Detection;120:103733.

[73] Asmare MH, Woldehanna F, Janssens L, Vanrumste B. Rheumatic Heart Disease Detection Using Deep Learning from Spectro-Temporal Representation of Un-segmented Heart Sounds. In: 2020 42nd Annual International Conference of the IEEE Engineering in Medicine & Biology Society (EMBC). IEEE;. p. 168–71.

[74] Ali F, Hasan B, Ahmad H, Hoodbhoy Z, Bhuriwala Z, Hanif M, et al. Detection of Subclinical Rheumatic Heart Disease in Children Using a Deep Learning Algorithm on Digital Stethoscope: A Study Protocol;11(8):e044070.

[75] Kapur G, Chen L, Xu Y, Cashen K, Clark J, Feng X, et al. Noninvasive Determination of Blood Pressure by Heart Sound Analysis Compared With Intra-Arterial Monitoring in Critically Ill Children—A Pilot Study of a Novel Approach:;20(9):809–16.

[76] Wang M, Guo B, Hu Y, Zhao Z, Liu C, Tang H. Transfer Learning Models for Detecting Six Categories of Phonocardiogram Recordings;9(3):86.

[77] Dong F, Qian K, Ren Z, Baird A, Li X, Dai Z, et al. Machine listening for heart status monitoring: Introducing and benchmarking hss—the heart sounds shenzhen corpus. IEEE journal of biomedical and health informatics. 2019;24(7):2082–92.

[78] Bentley P, Nordehn G, Coimbra M, Mannor S. The PASCAL Classifying Heart Sounds Challenge 2011 (CHSC2011) Results;. http://www.peterjbentley.com/heartchallenge/index.html.

[79] Liu C, Springer D, Moody B, Silva I, Johnson A, Samieinasab M, et al. Classification of heart sound recordings-the physionet computing in cardiology challenge 2016. PhysioNet. 2016.

[80] Yaseen, Son GY, Kwon S. Classification of Heart Sound Signal Using Multiple Features;8(12):2344.

[81] Kazemnejad A, Gordany P, Sameni R. EPHNOGRAM: A Simultaneous Electrocardiogram and Phonocardiogram Database. PhysioNet. 2021.

[82] Sameni R, Samieinasab M. Shiraz university fetal heart Sounds database”(version 1.0.1). PhysioNet. 2021.

[83] Winther S, Nissen L, Schmidt SE, Westra J, Andersen IT, Nyegaard M, et al. Advanced Heart Sound Analysis as a New Prognostic Marker in Stable Coronary Artery Disease;2(2):279–89.

[84] Humayun AI, Ghaffarzadegan S, Ansari MI, Feng Z, Hasan T. Towards Domain Invariant Heart Sound Abnormality Detection Using Learnable Filterbanks;24(8):2189–98.

[85] Wang P, Fan E, Wang P. Comparative analysis of image classification algorithms based on traditional machine learning and deep learning. Pattern Recognition Letters. 2021;141:61–7.

[86] Nassif AB, Shahin I, Attili I, Azzeh M, Shaalan K. Speech recognition using deep neural networks: A systematic review. IEEE access. 2019;7:19143–65.

[87] Otter DW, Medina JR, Kalita JK. A survey of the usages of deep learning for natural language processing. IEEE transactions on neural networks and learning systems. 2020;32(2):604–24.

[88] Oh SL, Hagiwara Y, Raghavendra U, Yuvaraj R, Arunkumar N, Murugappan M, et al. A deep learning approach for Parkinson’s disease diagnosis from EEG signals. Neural Computing and Applications. 2020;32:10927–33.

[89] Jamshidi M, Lalbakhsh A, Talla J, Peroutka Z, Hadjilooei F, Lalbakhsh P, et al. Artificial intelligence and COVID-19: deep learning approaches for diagnosis and treatment. Ieee Access. 2020;8:109581–95.

[90] Wolf T, Debut L, Sanh V, Chaumond J, Delangue C, Moi A, et al. Transformers: State-of-the-art natural language processing. In: Proceedings of the 2020 conference on empirical methods in natural language processing: system demonstrations; 2020. p. 38–45.

[91] Gong Y, Chung YA, Glass J. Ast: Audio spectrogram transformer. arXiv preprint arXiv:210401778. 2021.

[92] Lin T, Wang Y, Liu X, Qiu X. A survey of transformers. AI Open. 2022.

[93] Niu Z, Zhong G, Yu H. A review on the attention mechanism of deep learning. Neurocomputing. 2021;452:48–62.

[94] Clifford GD, Liu C, Moody B, Springer D, Silva I, Li Q, et al. Classification of normal/abnormal heart sound recordings: The PhysioNet/Computing in Cardiology Challenge 2016. In: 2016 Computing in cardiology conference (CinC). IEEE; 2016. p. 609–12.

[95] Nkomo VT, Gardin JM, Skelton TN, Gottdiener JS, Scott CG, Enriquez-Sarano M. Burden of Valvular Heart Diseases: A Population-Based Study;368(9540):1005–11.

[96] Vahanian A, Beyersdorf F, Praz F, Milojevic M, Baldus S, Bauersachs J, et al. 2021 ESC/EACTS Guidelines for the Management of Valvular Heart Disease:1–72.

[97] Otto CM, Nishimura RA, Bonow RO, Carabello BA, Erwin JP, Gentile F, et al. 2020 ACC/AHA Guideline for the Management of Patients With Valvular Heart Disease;77(4):e25–e197.

[98] Gardezi SK, Myerson SG, Chambers J, Coffey S, d’ Arcy J, Hobbs FR, et al. Cardiac Auscultation Poorly Predicts the Presence of Valvular Heart Disease in Asymptomatic Primary Care Patients;104(22):1832–5.

[99] Abbas Q, Hussain A, Baig AR. Automatic Detection and Classification of Cardiovascular Disorders Using Phonocardiogram and Convolutional Vision Transformers. Diagnostics. 2022;12(12):3109.

[100] Van Der Linde D, Konings EE, Slager MA, Witsenburg M, Helbing WA, Takkenberg JJ, et al. Birth Prevalence of Congenital Heart Disease Worldwide: A Systematic Review and Meta-Analysis;58(21):2241–7.

[101] Hinton RB, Ware SM. Heart Failure in Pediatric Patients with Congenital Heart Disease;120(6):978–94.

[102] D’Alto M, Mahadevan VS. Pulmonary Arterial Hypertension Associated with Congenital Heart Disease;21(126):328–37.

[103] Burchill LJ, Huang J, Tretter JT, Khan AM, Crean AM, Veldtman GR, et al. Noninvasive Imaging in Adult Congenital Heart Disease;120(6):995–1014.

[104] Brown KL, Ridout DA, Hoskote A, Verhulst L, Ricci M, Bull C. Delayed Diagnosis of Congenital Heart Disease Worsens Preoperative Condition and Outcome of Surgery in Neonates;92(9):1298–302.

[105] Martin N, Lilly LS. The Cardiac Cycle: Mechanisms of Heart Sounds and Murmurs:29–45.

[106] Kumar K, Thompson WR. Evaluation of Cardiac Auscultation Skills in Pediatric Residents;52(1):66–73.

[107] Huang PK, Yang MC, Wang ZX, Huang YJ, Lin WC, Pan CL, et al. Augmented detection of septal defects using advanced optical coherence tomography network-processed phonocardiogram. Frontiers in Cardiovascular Medicine. 2022;9:1041082.

[108] Van Oort A, Blanc-Botden L, De Boo T, Van Der Werf T, Rohmer J, Daniels O. The Vibratory Innocent Heart Murmur in Schoolchildren: Difference in Auscultatory Findings between School Medical Officers and a Pediatric Cardiologist;15(6):282–7.

[109] Savarese G, Lund LH. Global Public Health Burden of Heart Failure;3(1):7.

[110] Goldberg LR, Jessup M. Stage B Heart Failure: Management of Asymptomatic Left Ventricular Systolic Dysfunction;113(24):2851–60.

[111] McDonagh TA, Metra M, Adamo M, Gardner RS, Baumbach A, Böhm M, et al. 2021 ESC Guidelines for the Diagnosis and Treatment of Acute and Chronic Heart Failure;42(36):3599–726.

[112] Heidenreich PA, Bozkurt B, Aguilar D, Allen LA, Byun JJ, Colvin MM, et al. 2022 AHA/ACC/HFSA Guideline for the Management of Heart Failure: A Report of the American College of Cardiology/American Heart Association Joint Committee on Clinical Practice Guidelines;145(18).

[113] Hofmann S, Groß V, Dominik A. Recognition of Abnormalities in Phonocardiograms for Computer-Assisted Diagnosis of Heart Failures. IEEE;. p. 561–4.

[114] Bauersachs R, Zeymer U, Brière JB, Marre C, Bowrin K, Huelsebeck M. Burden of Coronary Artery Disease and Peripheral Artery Disease: A Literature Review;2019.

[115] Ghadrdoost B, Haghjoo M, Firouzi A. Accuracy of Cardiogoniometry Compared with Electrocardiography in the Diagnosis of Coronary Artery Disease;4(1):1.

[116] Padmanabhan V, Semmlow JL. Dynamical Analysis of Diastolic Heart Sounds Associated with Coronary Artery Disease;22(3):264–71.

[117] Akay M. Harmonic Decomposition of Diastolic Heart Sounds Associated with Coronary Artery Disease;41(1):79–90.

[118] Akay M, Akay YM, Gauthier D, Paden RG, Pavlicek W, Fortuin FD, et al. Dynamics of Diastolic Sounds Caused by Partially Occluded Coronary Arteries;56(2):513–7.

[119] Pathak A, Mandana K, Saha G. Ensembled Transfer Learning and Multiple Kernel Learning for Phonocardiogram Based Atherosclerotic Coronary Artery Disease Detection;26(6):2804–13.

[120] Roberts K, Colquhoun S, Steer A, Reményi B, Carapetis J. Screening for Rheumatic Heart Disease: Current Approaches and Controversies;10(1):49–58.

[121] Reményi B, Wilson N, Steer A, Ferreira B, Kado J, Kumar K, et al. World Heart Federation Criteria for Echocardiographic Diagnosis of Rheumatic Heart Disease—an Evidence-Based Guideline;9(5):297–309.

[122] Manji RA, Witt J, Tappia PS, Jung Y, Menkis AH, Ramjiawan B. Cost–Effectiveness Analysis of Rheumatic Heart Disease Prevention Strategies;13(6):715–24.

[123] Smulyan H, Safar ME. Blood Pressure Measurement: Retrospective and Prospective Views;24(6):628–34.

[124] Zhang XY, Zhang YT. Model-Based Analysis of Effects of Systolic Blood Pressure on Frequency Characteristics of the Second Heart Sound. IEEE;. p. 2888–91.

[125] Bombardini T, Gemignani V, Bianchini E, Venneri L, Petersen C, Pasanisi E, et al. Arterial Pressure Changes Monitoring with a New Precordial Noninvasive Sensor;6(1):1–11.

[126] Bartels A, Harder D. Non-Invasive Determination of Systolic Blood Pressure by Heart Sound Pattern Analysis;13(3):249.

[127] Benza RL, Miller DP, Gomberg-Maitland M, Frantz RP, Foreman AJ, Coffey CS, et al. Predicting Survival in Pulmonary Arterial Hypertension: Insights from the Registry to Evaluate Early and Long-Term Pulmonary Arterial Hypertension Disease Management (REVEAL);122(2):164–72.

[128] Humbert M, Sitbon O, Chaouat A, Bertocchi M, Habib G, Gressin V, et al. Pulmonary Arterial Hypertension in France: Results from a National Registry;173(9):1023–30.

[129] Luo H, Lamata P, Bazin S, Bautista T, Barclay N, Shahmohammadi M, et al. Smartphone as an electronic stethoscope: factors influencing heart sound quality. European Heart Journal-Digital Health. 2022;3(3):473–80.

[130] Yan W, Noel G, Loke G, Meiklejohn E, Khudiyev T, Marion J, et al. Single fibre enables acoustic fabrics via nanometre-scale vibrations. Nature. 2022;603(7902):616–23.

[131] Li P, Hu Y, Liu ZP. Prediction of cardiovascular diseases by integrating multi-modal features with machine learning methods. Biomedical Signal Processing and Control. 2021;66:102474.

